# Retrospective development and evaluation of prognostic models for exacerbation event prediction in patients with Chronic Obstructive Pulmonary Disease using data self-reported to a digital health application

**DOI:** 10.1101/2020.11.30.20237727

**Authors:** F. P. Chmiel, D. K. Burns, J. B. Pickering, Alison Blythin, T. M. A. Wilkinson, M. J. Boniface

**Author notes:** These authors contributed equally to this work (T. M. A. Wilkinson, M. J. Boniface).

## Abstract

**Background:** Self-reporting digital applications provide a way of remotely monitoring and managing patients with chronic conditions in the community. Leveraging the data collected by these applications in prognostic models could provide increased personalisation of care and reduce the burden of care for people who live with chronic conditions. This study evaluated the predictive ability of prognostic models for prediction of acute exacerbation events in people with Chronic Obstructive Pulmonary Disease using data self-reported to a digital health application.

**Methods:** Retrospective study evaluating the use of symptom and Chronic Obstructive Pulmonary Disease assessment test data self-reported to a digital health application (myCOPD) in predicting acute exacerbation events. We include data from 2,374 patients who made a total of 68,139 self-reports. We evaluated the degree to which the different variables self-reported to the application are predictive of exacerbation events and developed both heuristic and machine-learnt models to predict whether the patient will report an exacerbation event within three days of self-reporting to the application. The model’s predictive ability was evaluated on self-reports from an independent set of patients.

**Findings:** Users self-reported symptoms and standard Chronic Obstructive Pulmonary Disease assessment tests display correlation with future exacerbation events. Both a baseline model (AUROC 0.655 (95 % CI: 0.689-0.676)) and a machine-learnt model (AUROC 0.727 (95 % CI: 0.720-0.735)) showed moderate ability in predicting exacerbation events occurring within three days of a given self-report. While the baseline model obtained a fixed sensitivity and specificity of 0.551 (95 % CI: 0.508-0.596) and 0.759 (95 % CI: 0.752-0.767) respectively, the sensitivity and specificity of the machine-learnt model can be tuned by dichotomizing the continuous predictions it provides with different thresholds.

**Interpretation:** Data self-reported to healthcare applications designed to remotely monitor patients with Chronic Obstructive Pulmonary Disease can be used to predict acute exacerbation events with moderate performance. This could increase personalisation of care by allowing pre-emptive action to be taken to mitigate the risk of future exacerbation events. It is plausible future studies could improve the accuracy of these models by either the inclusion of symptom information recorded with greater granularity or including variables not considered in our study, for example vital signs, information on activity, local environmental data, and lifestyle information.

**Funding:** This project was funded by the European Union’s Horizon 2020 research and innovation programme under grant agreement No 780495 BigMedilytics.

## Research in context

### Evidence before this study

Avoiding exacerbations is desirable for COPD patients because it likely leads to both a higher quality of and longer life. Several studies have attempted to risk-stratify patients according to their exacerbation risk. Generally, these prognostic models predict a patient’s exacerbation frequency to guide long-term care, as opposed to predicting acute events to provide advanced warning of and the opportunity to mitigate the risk of symptom exacerbation. Prediction of patients exacerbation frequency is important since it allows more informed decisions about care, for example in the decision to prescribe pharmacological interventions (such as inhaled corticosteroids or phosphodiesterase-4 inhibitors) which reduce exacerbation frequency but are strongly associated with harm. However, the accurate prediction of acute events is also of significant utility since it would enable pre-emptive treatment strategies (as opposed to reactive care) which could prevent exacerbations and reduce hospitalisation.

### Added value of this study

We demonstrated that data self-reported to a digital health application can be used to predict short-term future exacerbation events. This outcome is distinct to the majority of studies, since we demonstrate it is likely acute exacerbation events can be predicted several days in advance using data recorded by a digital health application as opposed to simply the predicting a patient’s yearly exacerbation frequency.

### Implications of all the available evidence

Our study could allow the implementation of new exacerbation mitigation strategies aimed to avoid exacerbation events in the short-term. Increased level of care personalization will be facilitated by the integration of prognostic models into clinically validated digital applications for managing COPD patients. Increasing the number of variables available to these models would likely improve their discriminative ability further.

## Introduction

Chronic Obstructive Pulmonary Disease (COPD) is a collection of progressive lung diseases, characterised by breathing difficulties and an irreversible reduction of lung function. It is one of the most prevalent chronic conditions in the world (in England 2.19 % of the population are expected to have a confirmed COPD diagnosis by 2030^3^) and, the absence of a cure, means its represents a significant burden for patients who have to manage the condition on a daily basis^1,2^. A key characteristic of managing COPD is in mitigating the risk of ‘exacerbation events’, which can be defined as an acute, sustained worsening of a patient’s condition that necessitates a change in medication^4^. Exacerbations accelerate lung function decline and evidence suggests that the frequency of exacerbations increases with decreasing lung function^5–7^. Minimizing the number of exacerbation events can therefore have a significant impact on the prognosis for COPD patients. Currently, several methods exist to help control exacerbation events, including pharmacological interventions, pulmonary rehabilitation, and self-management programmes^8^. There is also an identified clinical need to predict exacerbation events in advance to personalize COPD treatment and offer the opportunity to provide targetted pre-emptive interventions^9,10^.

In recent years, the advent of mobile health applications has facilitated increased remote management and care of patients with COPD^11,12^. These applications allow the recording of temporally dense information about a patient’s condition which allow (near) real-time monitoring of a patient’s symptoms, providing clinicians with a source of data to help them understand how the patient is managing their condition and gain an insight into the patient’s exacerbation frequency and severity. For the patient, digital health applications provide both an access point for educational content about their condition and the opportunity to improve their self-care, leading to better long-term management^13^. In the context of COPD, there is an opportunity to increase the efficacy of digital health applications further, by leveraging the data they collect to predict acute exacerbation events and provide personalized alerts to the patient. These alerts could facilitate the following of a clinically validated, personalized intervention programme in an attempt to mitigate the occurrence of an acute exacerbation event.

In this report we present a retrospective study making use of data collected by the myCOPD mobile application, an NHS approved, clinically validated application for persons with a diagnosis of COPD^13^. The application assists with the management of COPD by providing educational content alongside a digital, momentarily assessed symptom diary. Using the application, users self-report on the COPD-related symptoms they are currently experiencing, as well as information which characterises their long-term COPD status (i.e., the COPD Assessment Test). Using statistical analysis and machine-learning methods we evaluate the effectiveness of exploiting this simple, self-reported information to predict exacerbation events in the near-future and discuss how such predictions could be used to improve the long-term outcomes for people with COPD.

## Methods

### Dataset description

We used an anonymised extract of daily user self-reports submitted to the myCOPD application between 01/01/2017 and 31/12/2019 (inclusive). A single report features a self-assessed symptom score (Figure 1) which is a four-point scale ranging from normal symptoms to a severe deterioration of symptoms requiring hospitalisation (we encode this as an ordinal variable between 1 and 4 indicating increasing severity of COPD related symptoms, see Figure 1 b). Users also perform a COPD Assessment Test (CAT) at regular (approximately monthly) intervals. The CAT is an eight question assessment and yields a ‘score’ between 0 and 40, where higher values indicate a more severe impact of COPD on a user’s overall health^14^. It is validated and is an accepted way of quantifying the burden of COPD on someone’s life^15,16^. In addition to these scores, our dataset also features additional demographic and lifestyle information self-reported to the application. These include patient age, gender, current smoking status, and the number of years they have been smoking for.

**Figure 1.**
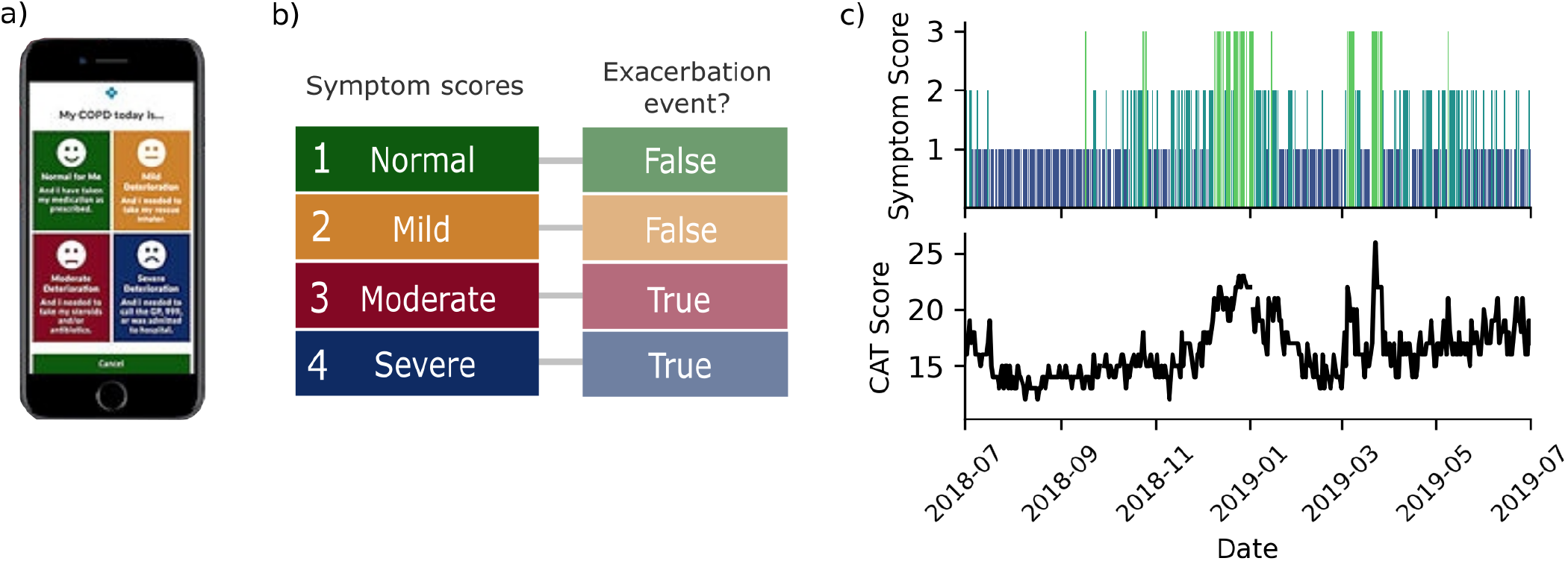
Self-reported symptom scores and COPD Assessment Test (CAT) results. Landing page of the myCOPD smartphone application where users self-report their daily symptom score. b) Symptom score rankings and classification of whether this score corresponds to an exacerbation event, as defined in the context of this work. c) Example user (with high reporting frequency) self-reporting timeline where the top panel displays self-reported symptom scores and the bottom panel self-reported CAT results.

### Defining exacerbation events

We use a symptom-based definition for exacerbation events where an event is marked to have occurred if a patient self-reports either the use of medication to control their symptoms or hospitalization resulting from their condition, corresponding to a self-reported symptom score of 3 or 4 respectively (Figure 1)^17^.

### Cohort selection and dataset segregation

In total 5,170 users were included in the extract who reported a total number of 94,882 reports in the study period. User registration was incremental (i.e., not all users registered at the same time) throughout the period of the study and self-reports were not necessarily submitted every day. To create our study cohort we followed a selection process outlined in Figure 2. First, isolated self-reports (those in which a second report was not made within 3 days) were removed because the target variable could not be reliabilty calculated (see ‘Predicting exacerbation events’). Next, reports from anomalous users (those only reporting exacerbation events or entering self-reports before their registration date) were removed. After removal of these reports we obtained our final study cohort, featuring 68,139 reports from 2,374 unique users.

**Figure 2.**
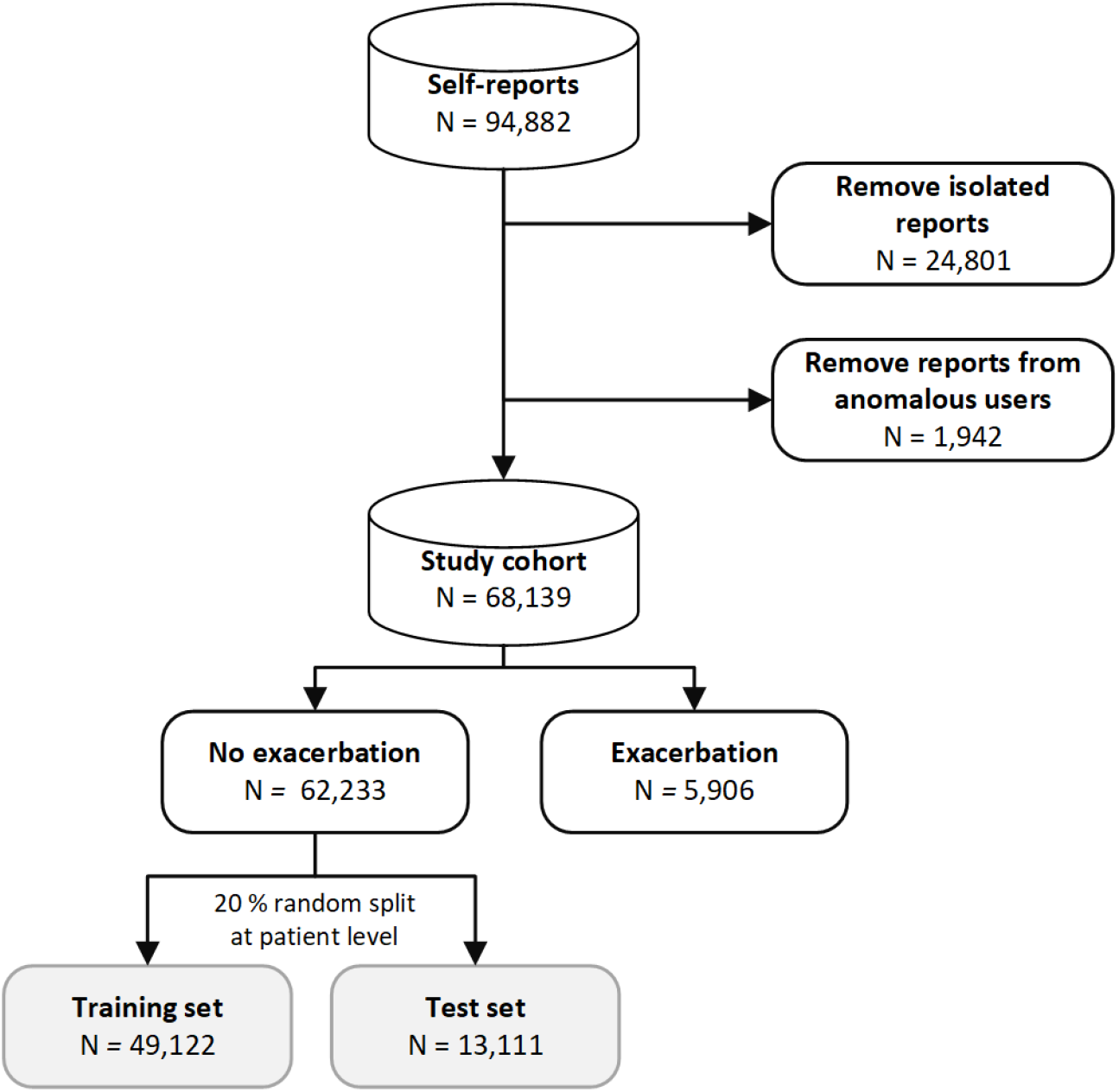
Selection of self-reports in our study cohort. Isolated reports (N=24,801) are those without a subsequent report in the following three days. Anomalous users (N=1,942) are those who only report exacerbation events or self-report to the myCOPD application before their registration date.

### Predicting exacerbation events

For each daily self-report, we created a binary variable which indicated whether a report is followed by an exacerbation event in the following three days. A three day window was chosen to be close enough to the future exacerbation event for any signal to be present in the data but sufficiently far from the event such that a range of pre-emptive actions could be available to patients. For the development of the prognostic models, we selected only reports in which the patient did not report an exacerbation event on the same day (N=49,122, Figure 2). We then randomly assigned all reports from 20 % of patients to a hold-out test set and the remaining to a training set. We created a baseline, heuristic model which uses a users most recently reported symptom score, assigning users a either a low risk (1.7 %) or heightened risk (7.2 %) of future exacerbation, where percentages (brackets) correspond to the mean three-day exacerbation rate for all reports in the training set with symptom scores of one or two respectively.

Our supervised machine-learning models made use of patient demographics, lifestyle information, self-reported information, and aggregate features which summarizes a patients (recent) self-reporting history. A full schema of variables used by our models is presented in Supplementary Table 1. For our predictive models we used logistic regression and a Random Forest classifier each trained by 5-fold, grouped cross-validation at the user level, i.e., reports from a single user appear exclusively in either the training or validation fold. Missing CAT scores were forward-filled imputed at the user level where possible. All other missing values were filled using mean imputation within fold. Either target or ordinal encoding was used for all categorical variables (Supplementary Table 1). Model hyperparameters were optimized on the out-of-fold validation samples using Bayesian optimization via the Tree Parzen Estimator algorithm as implemented in the hyperopt Python library^18,19^. Model performance was evaluated on the hold-out test set and 95 % confidence intervals were estimated using bootstrapping. To create a binary decision of exacerbation risk, model predictions were dichotimized with thresholds chosen to yield either a fixed specificity or the maximum Youden’s J statistic on the test set^20^.

## Results

Our cohort self-reporting to the application featured 2,374 users in total, of which most (70.4 %) were between the ages of 60 and 79, inclusive (Table 1). Only 27.3 % of users reported their gender, with 419 males reporting compared to 231 females. A large fraction (49.7 %) of users reported their smoking history, with 86.5 % of those reporting being either a current or ex-smoker (Table 1). Out of the self-reports included in this study patients reported 5,906 self-reports which correspond to an exacerbation event in the context of this work, 8.7 % of total reports (Figure 2).

**Table 1.** Patient demographics and smoking status in our cohort. All information was self-reported to the myCOPD application. Patient information relates to the time at which the patient first reported to the application (e.g., if there smoking status changed, this table summarizes the first reported status). There are 2,374 unique patients in total.

|  | Patient count (%) |
| --- | --- |
| **Age group** |  |
| Missing | 10 (0.4) |
| 19-29 | 7 (0.3) |
| 30-39 | 39 (1.6) |
| 40-49 | 89 (3.7) |
| 50-59 | 325 (13.7) |
| 60-69 | 791 (33.3) |
| 70-79 | 881 (37.1) |
| 80-89 | 212 (8.9) |
| 90-99 | 15 (0.6) |
| 100-110 | 5 (0.2) |
| **Gender** |  |
| Missing | 1,724 (72.6) |
| Female | 231 (9.7) |
| Male | 419 (17.6) |
| **Smoking status** |  |
| Missing | 1,217 (51.3) |
| Ex-smoker | 843 (35.3) |
| Non-smoker | 156 (6.6) |
| Smoker | 158 (6.7) |

In Figure 3 we investigate the two metrics self-reported to the myCOPD application and their relation to self-reported exacerbation events. Panels a to d of Figure 3 display the correspondence between between symptom scores and the CAT results when self-reported on the same day. While for each symptom score users nearly report the full range of CAT scores, there is a clear correlation between CAT scores and symptom scores, with users reporting higher symptom scores more likely to also report a higher CAT score. For example, the mean CAT score reported when a user reports a symptom score of one is 13.5, which is significantly lower (p<0.001) than the mean CAT score (19.5) when a user reports a symptom score of two. Such a correlation is to be expected, research has shown increased CAT scores correlate with an increased exacerbation frequency^21^, which in turn would lead to higher symptom scores being reported in our study.

**Figure 3.**
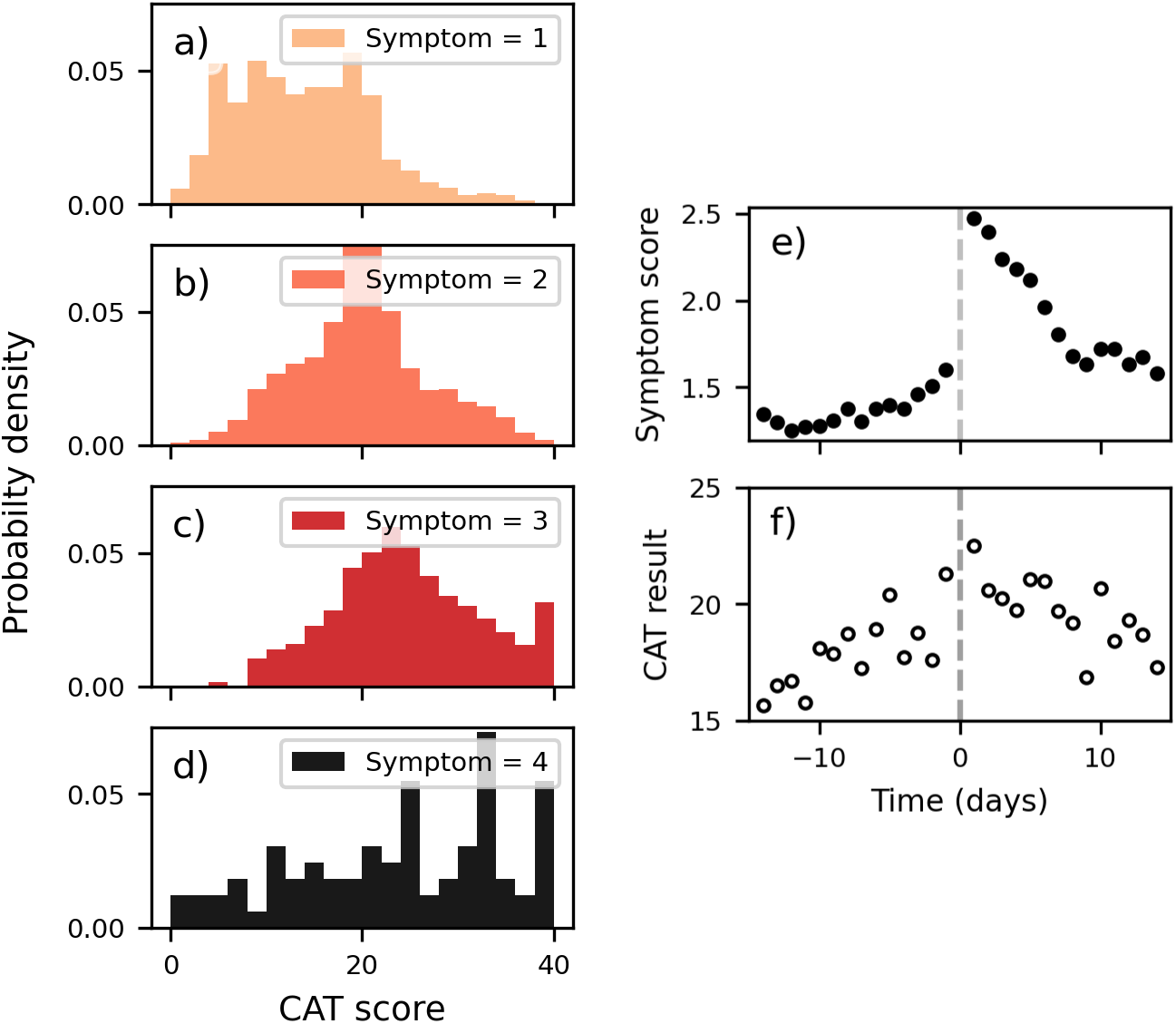
Self-reported symptom scores and results of COPD Assessement Tests for reports in our cohort. a-d) Displays the self-reported CAT result stratified by the self-reported symptom score (row) on the day of test completion. e) Mean self-reported symptom scores in the days preceding (and following) a day where a patient self-reports their first exacerbation event. f) Mean self-reported result of CAT in the days preceding (and following) a day where a patient self-reports their first exacerbation event. Grey dashed lines in all panels highlight the day of the first reported exacerbation event (time = 0 days). Panels e and f indicate that exacerbation events can be associated with a worsening of symptom scores and CAT results several days in advance of the event. The width of the observed ‘peaks’ (see panel e, right of dashed line) following the start of the exacerbation event demonstrate that exacerbation events can be multiple day events.

In Figure 3 e and f, we evalute the sensitivity of symptom scores and CAT results to future exacerbation events. We display how the mean of the two reported variables change in days preceeding (and following) the day in which users self-report their first exacerbation event to the application (not necessarily their first ever exacerbation event). By inspection of Figure 3 e we see that the mean reported symptom score in proximity of the first reported exacerbation event increases, indicating that in the days preceding their first exacerbation event users are increasingly likely to report a mild deterioration of symptoms (symptom score of 2). Changes in the mean symptom score calculated across all users can be seen several days in advance, suggesting that at least some users are observing a mild deterioation in symptions several days in advance of exacerbation events. For days subsequent to users first self-reported exacerbation (right of the dashed line in Figure 3 e) the mean symptom score initially exceeds 2, showing that self-reported exacerbation events can be multi-day events - consistent with current understanding of exacerbation events^4^. Similarily, in Figure 3 f the reported mean CAT result is observed to increase in magnitude in the days preceeding an exacerbation event and then decrease (at a slower rate) following a reported event. Overall, these trends indicate there is potential in using these self-reported variables to predict at least a subset of exacerbation events in advance.

Next we discuss the baseline prognostic model, the model captures the key feature about exacerbation events observed in our data: persons reporting a mild deterioration of symptoms are significantly (p<0.001) more likely to experience an exacerbation event in the next three days compared to those reporting normal symptoms, with a relative risk of 4.16 (95 % CI: 3.8-4.5). On the hold-out test set the baseline model obtained an AUROC of 0.655 (95 % CI: 0.689-0.676). In Figure 4, we display the Receiver Operating Characteristic (ROC) curve for our baseline model alongside the two machine-learnt models, a logistic regression model (solid grey line) which does not consider variable interactions and a random forest classifier (solid black line) which does. The logistic regression model obtained an AUROC of 0.697 (95 % CI: 0.689-0.711) and the Random Forest model 0.727 (95 % CI: 0.720-0.735), on the hold-out test (Table 2). The significantly higher (p<0.001) performance of the Random Forest model suggests either interactions between variables are important in discriminating between reports associated with exacerbation within three days or non-linear relations are present.

**Table 2.**
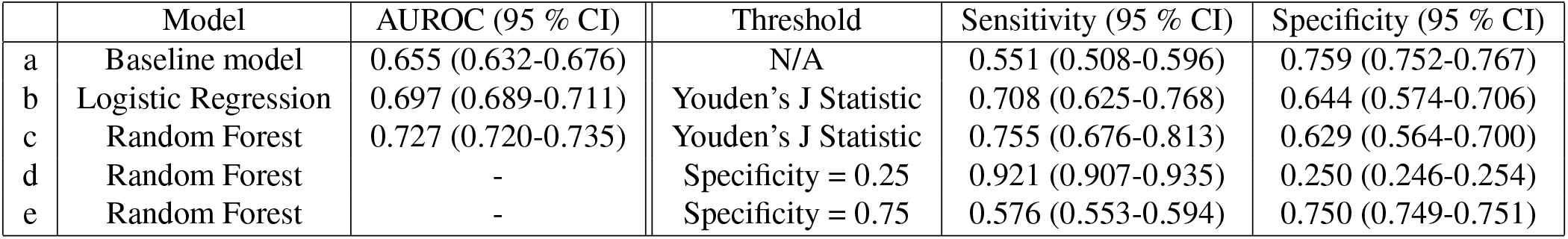
Model performances evaluated on the hold-out test set. The AUROC column denotes the area under the receiving operator curve (Figure 4) for each model. The three right-most columns display the sensitivity and specifivity of models at predicting exacerbations with different thresholds used to dichotmize the predictions. The baseline model is already binary and only has one non-trivial configuration but the threshold used to dichtomize the machine learning models (b-e) can be tuned to suit the intended context of the model. The maximum of Youden’s J stastic is used as a baseline criterion for dichtomizing the prediction (model b and c) and other cut-offs yielding fixed specificities are investigated for the Random Forest model.

|  | Model | AUROC (95 % CI) | Threshold | Sensitivity (95 % CI) | Specificity (95 % CI) |
| --- | --- | --- | --- | --- | --- |
| a | Baseline model | 0.655 (0.632-0.676) | N/A | 0.551 (0.508-0.596) | 0.759 (0.752-0.767) |
| b | Logistic Regression | 0.697 (0.689-0.711) | Youden's J Statistic | 0.708 (0.625-0.768) | 0.644 (0.574-0.706) |
| c | Random Forest | 0.727 (0.720-0.735) | Youden's J Statistic | 0.755 (0.676-0.813) | 0.629 (0.564-0.700) |
| d | Random Forest | - | Specificity = 0.25 | 0.921 (0.907-0.935) | 0.250 (0.246-0.254) |
| e | Random Forest | - | Specificity = 0.75 | 0.576 (0.553-0.594) | 0.750 (0.749-0.751) |

**Figure 4.**
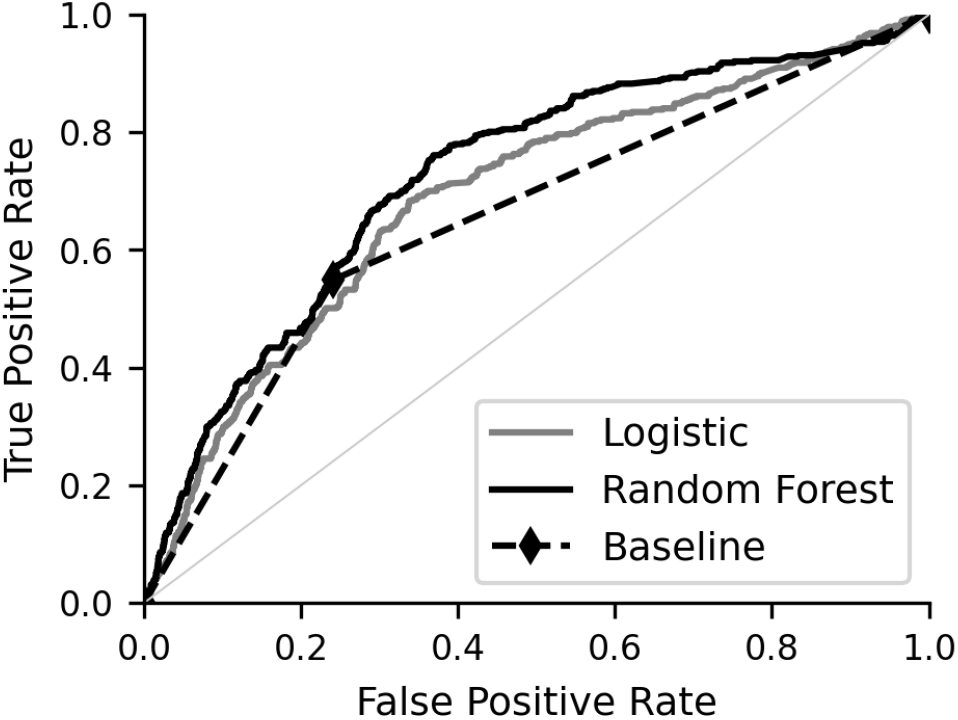
Receiver operating characteristic curve of our models evaluated on the patient hold-out test set. The baseline model (dashed line) has only one, non-trivial threshold for dichotimizing the prediction (diamond marker), where as the machine learnt models have a large number of thresholds which needs to be optimized to suit the use-case (so called sensitivity-specifitity trade-off).

In Table 2 we present the sensitivity and specificity of the baseline model and the machine-learnt models evaluted on the hold-out test set. While the baseline model is already dichotomized, for the machine-learnt models a threshold must be chosen to binarize the continuous exacerbation risks they produce. The baseline model obtained a sensitivity of 0.551 (95 % CI: 0.508-0.596) with specificity of 0.759 (95 % CI: 0.752-0.767). While neither machine-learnt model significantly outperform the baseline model at the same specificity (e.g., compare model a and e in Table 2), the tuning of the threshold used to dichotomize the machine-learnt models predictions can lead to a range of sensitivity and specificities (compare models c, d and e in Table 2) on the hold-out test which could be tuned to match different escalation policies and interventional strategies. For example, the random forest model can be tuned to yield a sensitivity of 0.921 (95 % CI: 0.907-0.935) or 0.576 (95 % CI: 0.553-0.594) with respective sensitivities of 0.250 (95 % CI: 0.246-0.254) or 0.750 (95 % CI: 0.749-0.751).

## Discussion

Symptom information self-reported to the myCOPD application displayed correlation to the start of future exacerbation events (Figure 2 e) and we found machine-learnt models utilizing this and other sources of self-reported data were able to identify patients at risk of exacerbation within three days with moderate discriminative ability (AUROC 0.727 (95 % CI: 0.720-0.735)).

Our analysis has shown strong evidence that users self-report changes in symptoms prior to exacerbation events which could be leveraged to predict future events. Machine-learnt models are well positioned to take advantage of these trends because they can learn complex, non-linear relations between variables which are inaccessible to hand crafted models and, despite the complexity of the models, they can be easily integrated into digital applications. This would greatly facilitate the uptake of the model since they can be directly implemented in an active, digital ecosystem for patient care. Conversely, this is also a limitation of machine-learnt models deployed in such a fashion since they will likely generalize poorly outside the digital application for which they were designed.

The predictive performance of the machine-learnt models we trained are limited by the number of variables available within our dataset. There are many factors not considered by our model which have the potential to be predictive of acute COPD exacerbations. For example, various health and lifestyle factors (including comorbidity and socioeconomic status) not known to our model are believed to impact COPD exacerbation frequency^22^. These could be acquired from a patient’s medical record or collected with questionnaires and used by the algorithm to further refine its predictions. Additionally, since self-reported deterioration in symptoms can occur several days before an exacerbation (Figure 2 e), it is reasonable that symptom information could be collected in a more granular manner in the days preceeding an exacerbation event. This could be achieved with medical devices (e.g. smart inhalers), wearables designed to monitor person’s lung function and COPD related physiological and behavioural variables, or through more granular self-reporting of symptoms through the digital application^23^. While research has shown that environmental factors (such as pollution and weather) localised to a user could potentially be predictive of future COPD exacerbation events^24,25^, available data sets from pollution monitoring stations are limited by spatial resolution, as are proxies for exposure, such as distance of residence from roadside or other point sources. While such datasets may be appropriate for population-level analyses, they are not well suited for understanding effects of pollution on a heterogeneous population at the individual level. Furthermore, such approaches do not allow incorporation of individual movement, with error as soon as the individual moves away from the monitoring/residential site, and potentially significant errors relating to differential exposures in microenvironments (e.g. indoors at home, in vehicles). Further work is needed to study the chemical biological interaction between individual movement, hyperlocal pollution monitoring, and the prediction of acute COPD exacerbation.

Our study has several other limitations. Firstly, while our self-reports were collected in a prospective fashion using momentary assessments of patients COPD symptoms, reporting compliance and individual variation in reporting behaviours could still be determintal to our results, with exacerbation frequencies or severity being misreported^26^. Imperfect reporting compliance also acts to limit the amount of data available to our machine-learnt model which may limit their predictive ability. Reporting compliance could be improved by increased clinician engagement but it is unlikely clinicians will have sufficient time to monitor and engage with the patient self-reports to a sufficient level. Finally, we used a validation set featuring unique patients that were extracted from the same study population as those in the training set and therefore may not meet all requirements to be considered independent. A more robust validation set containing an external cohort of patients is desirable.

For the machine-learnt models it is important to match their configuration to the escalation policy. If models are used as a binary alert system, this is achieved by analysing the (so-called) sensitivity-specificity trade-off, considered in Table 2 or by inspection of Figure 4. In Table 2 three configurations of the Random Forest model are chosen (models c, d, and e) which yield different sentivities and specificites. For example, model d in Table 2 uses a threshold chosen to obtain a specificity of 0.25 on the test set and achieves a sensitivity of 0.921 (95 % CI: 0.907-0.935). This configuration could be appropiate if false positives are not of signficant concern (e.g., if the prescribed intervention plan is of little risk to the patient). Ultimately, different configurations allow flexiblity in the resulting escalation policies and is the key advantage of the machine-learnt models compared to the baseline model.

In order to influence clinical outcome beneficially, a predictive model needs to enable a pre-emptive intervention with the likely impact maximised the earlier this occurs. In the context of COPD exacerbations interventions may include increased use of inhaled medication or additional administration of rescue packs of oral antibiotics and corticosteroids^28^. The current standard of care has been for patients to take these treatments when they are actually exacerbating, which creates a reactive model of care requiring significant clinical deterioration to have occurred before an intervention is started. Our own work has shown that early treatment of exacerbations is associated with improved clinical outcomes including faster recovery times^27^. Therefore, if presented appropriately this risk prediction model could enable patients to self-manage more effectively intervening before life-threatening inflammation and infection can become established.

To conclude, data self-reported to a digital health application, designed for the management of people with COPD, can be used to identify users at risk of exacerbation within three days with moderate discriminative ability (AUROC 0.727 (95 % CI: 0.720-0.735)). Further research utilizing additional linked data (particularily from medical devices such as smart inhalers and environmental sensors) are expected to increase the accuracy of these models.

## Data Availability

Data will be made available upon reasonable request to persons with a university affliation. Requestors will need appropiate data protection, governance, and ethical review in place.

## Contributors

FPC performed the analysis with support from DKB and MJB. FPC wrote the first draft of the manuscript. FPC, JBP, MJB wrote the second draft of the manuscript. JBP obtained ethical and governance approvals. FPC, JBP, and MJB led the research project at UoS. AB managed the data extraction at mymhealth. TMAW and AB provided clinical insight. FPC, MJB and TMAW envisaged the research. All other authors contributed to future iterations of the manuscript.

## Acknowledgements

We acknowledge support and discussions with respect to concept and data analysis from Dr B. Arbab-Zavar and Dr Zoheir Sabeur.

## Ethical approval and data governance

This work received ethics approval from the University of Southampton’s Faculty of Engineering and Physical Science Research Ethics Committee (ERGO/FEPS/52137) and was reviewed by the University of Southampton Data Protection Impact Assessment panel (DPIA 0045), with the decision to support the research.

